# Dynamic near-term risk prediction of clinically significant immune-related adverse events using longitudinal electronic health records

**DOI:** 10.64898/2026.10.06.26364907

**Authors:** Liyuan Gong, Alice Agyekum, Nitish Aswani, Harry Reyes Nieva, Chin Hur

**Author notes:** Corresponding author: Chin Hur.

## Abstract

**Objective:** To develop and evaluate a dynamic model that repeatedly estimates the 30-day risk of a first grade ≥2 immune-related adverse event (irAE) during immune checkpoint inhibitor (ICI) therapy.

**Materials and Methods:** Adults initiating ICI therapy during 2015-2022 comprised the development cohort; 2023 initiators formed a nonoverlapping internal temporal evaluation cohort. Every 21 days during the first treatment year, models used available EHR information to predict a first structured phenotype-defined grade ≥2 irAE during the next 30 days. Evaluation included pooled and within-landmark discrimination, precision–recall performance, calibration, and patient-clustered bootstrap confidence intervals. Feature-state ablations and alternative update schedules examined the contribution of dynamic information.

**Results:** Development included 1,522 patients and 18,744 evaluable landmarks; temporal evaluation included 526 patients and 6,263 landmarks, of which 184 were positive (2.94%). The multiscale XGBoost model achieved pooled AUROC 0.716 (95% CI, 0.676-0.755), AUPRC 0.067, within-landmark AUROC 0.672 (95% CI, 0.631-0.716), Brier score 0.0281, and log loss 0.1245. The time-aware baseline achieved AUROC 0.631 and within-landmark AUROC 0.536. Updated treatment exposure and laboratory trajectories accounted for most of the gain. A complete-state extension achieved AUROC 0.719 and AUPRC 0.073. The 21-day schedule had the highest discrimination among tested cadences.

**Conclusion:** Longitudinal EHR updating improved near-term irAE prediction beyond pretreatment characteristics and treatment time. Phenotype adjudication and independent evaluation of calibration, transportability, and clinical utility are needed before implementation.

## Introduction

Immune checkpoint inhibitors (ICIs) have transformed cancer treatment but can trigger immune-related adverse events (irAEs) affecting nearly any organ system. Moderate or severe toxicities can require treatment interruption, systemic immunosuppression, hospitalization, or permanent ICI discontinuation.^1–3^ Their timing, organ involvement, and severity vary across patients and treatment phases, requiring repeated reassessment of toxicity alongside the benefits of continued therapy.^1,3,4^ Guidelines emphasize surveillance and prompt symptom evaluation, but routinely implemented tools for updating individual near-term toxicity risk remain limited.^1–3^

Reliable identification of clinically significant irAEs from electronic health record (EHR) data is challenging. Diagnosis codes may incompletely capture events and do not establish attribution to ICI therapy.^5^ Natural language processing and large language models, including irAE-GPT, identify toxicities from clinical narratives; agentic approaches additionally extract temporality, severity, attribution, and certainty while assisting human review.^5–8^ These systems characterize documented events rather than estimate the risk of a new event within a future prediction window. This distinction separates establishing whether and when toxicity occurred from forecasting toxicity before its onset. Structured phenotyping offers a complementary approach by combining temporally aligned exposure, diagnoses, laboratory findings, and immunosuppressive treatment, as illustrated in a recent preprint.^9^ However, suspected irAEs must be distinguished from infection, cancer progression, other treatment toxicities, and preexisting disease.^1–3^ Clinical validation is therefore essential, particularly because inaccurate onset dates can move events across prediction-window boundaries. Dynamic prediction requires outcome labels whose event status, severity, organ attribution, and onset are evaluated against chart review.

Many irAE prediction studies use pretreatment clinical characteristics, routine blood tests, autoantibodies, or imaging.^10–13^ Other studies identify associations with T-cell characteristics, early B-cell changes, germline variants, and intestinal microbiota.^14–17^ These biomarkers provide complementary biological information, although specialized immune profiling and microbiome assays are not consistently available in routine care. Baseline-only estimates also cannot incorporate changes arising from ongoing exposure, concomitant therapies, evolving laboratory abnormalities, symptoms, or healthcare utilization. Consequently, patients with similar pretreatment profiles may develop different risk trajectories as new clinical information accumulates.

On-treatment approaches have associated early B-cell changes with subsequent toxicity, used repeated electronic patient-reported symptoms to predict irAE presence and onset, and incorporated diagnoses and chronological medication information into EHR-based graph models.^15,18,19^ These studies support updated assessment while motivating evaluation of recurrent prediction over a standardized near-term horizon. Longitudinal EHR data offer a practical foundation because they capture evolving clinical information without requiring an additional research assay. A fixed prediction horizon makes the clinical question explicit: which event-free patients may experience clinically significant toxicity soon?

Dynamic prediction nevertheless introduces methodological challenges. Patients contribute correlated prediction opportunities, event frequency and risk-set composition change during follow-up, and observation ends after toxicity, death, or loss of follow-up. Landmark analysis defines standardized prediction times and event-free risk sets using only information available at each prediction time.^20^ Extensions accommodate competing events and longitudinal biomarker histories.^21,22^ Landmark construction alone, however, does not resolve within-patient correlation or incomplete follow-up. Pooled discrimination may also reflect differences between treatment phases rather than discrimination among patients within the same phase. Evaluation should therefore assess performance across prediction opportunities and within individual landmarks, estimating uncertainty at the patient level to preserve dependence among repeated observations.^23^

In this study, we developed and evaluated a structured EHR phenotype for the first grade 2 or higher irAE, targeting toxicities likely to prompt clinical evaluation or altered management. We then constructed a dynamic landmark framework that updated risk throughout the first year of ICI therapy and estimated event probability during the subsequent 30 days. We hypothesized that longitudinal information would improve discrimination and probabilistic accuracy compared with pretreatment risk factors and treatment time alone. We also evaluated alternative algorithms, update schedules, feature-group ablations, and sensitivity to follow-up and phenotype definitions.

## Materials and methods Study design and data source

We conducted a retrospective longitudinal cohort study using structured electronic health record (EHR) data from Columbia University Irving Medical Center/NewYork-Presbyterian. Data were organized into an Observational Medical Outcomes Partnership (OMOP)-compatible representation and included demographics, diagnoses, medication exposures, procedures, encounters, laboratory measurements, and immune checkpoint inhibitor administrations.^24^ Data were extracted on May 30, 2026. The study was approved by the institutional review board (protocol ACYY1210), with a waiver of informed consent for retrospective secondary analysis.

## Study population

Eligible patients were adults aged ≥18 years with cancer, at least 365 days of EHR observation before ICI initiation, and qualifying ICI exposures documented on at least two distinct dates. The index date was the first recorded qualifying ICI administration. ICI exposures were identified using RxNorm ingredient concepts and their descendant drug concepts, encompassing PD-1, PD-L1, and CTLA-4 inhibitors and combination regimens, including PD-1 plus LAG-3 blockade. Patients initiating ICI therapy during 2015-2022 formed the development cohort, whereas those initiating therapy during 2023 formed the internal temporal evaluation cohort. The cohorts were mutually exclusive, with no patient contributing to both. The qualifying ICI agents and ingredient-level concept sets are provided in Supplementary Table S1.

## Outcome definition and chart review

The outcome was the first structured phenotype–defined irAE compatible with grade ≥2 after ICI initiation. Candidate events were identified using organ-specific diagnoses, compatible laboratory abnormalities, and supporting clinical management, including new systemic corticosteroid or other immunosuppressive treatment and hormone replacement for endocrine toxicities. Corticosteroid anchors were restricted to oral or parenteral administration; topical, inhaled, and other local preparations were excluded. Diagnosis-only events required repeated documentation but did not independently establish grade ≥2 toxicity. Pre-existing conditions and treatments were assessed using the phenotype-specific baseline exclusion rules. Diagnosis codes and medication concept sets are provided in Supplementary Tables S2 and S3, respectively, with detailed anchoring criteria in Supplementary Methods.

The event date was the earliest date satisfying the qualifying phenotype criteria. At each landmark, the binary outcome indicated whether the first qualifying event occurred during the subsequent 30 days (days 1-30). Landmarks on or after that event were excluded. Grade 1 events were not outcomes, and patients with grade 1 toxicity alone remained at risk. ICI interruption or discontinuation alone did not establish an irAE or its severity. Structured labels represented grade ≥2-compatible events rather than adjudicated CTCAE grades.

Structured event dates were audited against the derived 30-day labels, with no date-logic mismatches identified. Manual chart review was completed for 296 patients sampled across five groups: 60 each from phenotype Rules 1, 3, and 4 and the phenotype-negative group (Rule 5), and 56 from Rule 2. Review involved L.G., A.A., and C.H. to assess clinical attribution, severity, and event timing. Chart-review methods and findings are reported in Supplementary Methods and Supplementary Table S4.

## Dynamic landmark design

Risk was updated every 21 days from treatment day 0 through day 357, with each landmark predicting the subsequent 30 days.^25^ This schedule approximated a 3-week ICI treatment cycle. Prediction windows overlapped, allowing a single event to generate up to two positive landmark observations. Repeated observations were addressed through patient-level data partitioning and bootstrap resampling. Patients were eligible at each landmark if they were alive, under observation, and free of a previous qualifying irAE. Predictors used only information available at or before the landmark. Follow-up ended at death or last follow-up. A first qualifying irAE occurring within the prediction window and before death was assigned an outcome of 1. Death within the window without a preceding qualifying irAE was treated as a competing event and assigned an outcome of 0, as was remaining event-free through day 30. Landmarks with follow-up ending before day 30 without either a qualifying irAE or a documented death were excluded because outcome ascertainment was incomplete. Sensitivity analyses regenerated the feature pipeline using 14-and 30-day updating intervals while retaining a 30-day prediction horizon and fixed hyperparameters (Figure 1).

**Figure 1.**
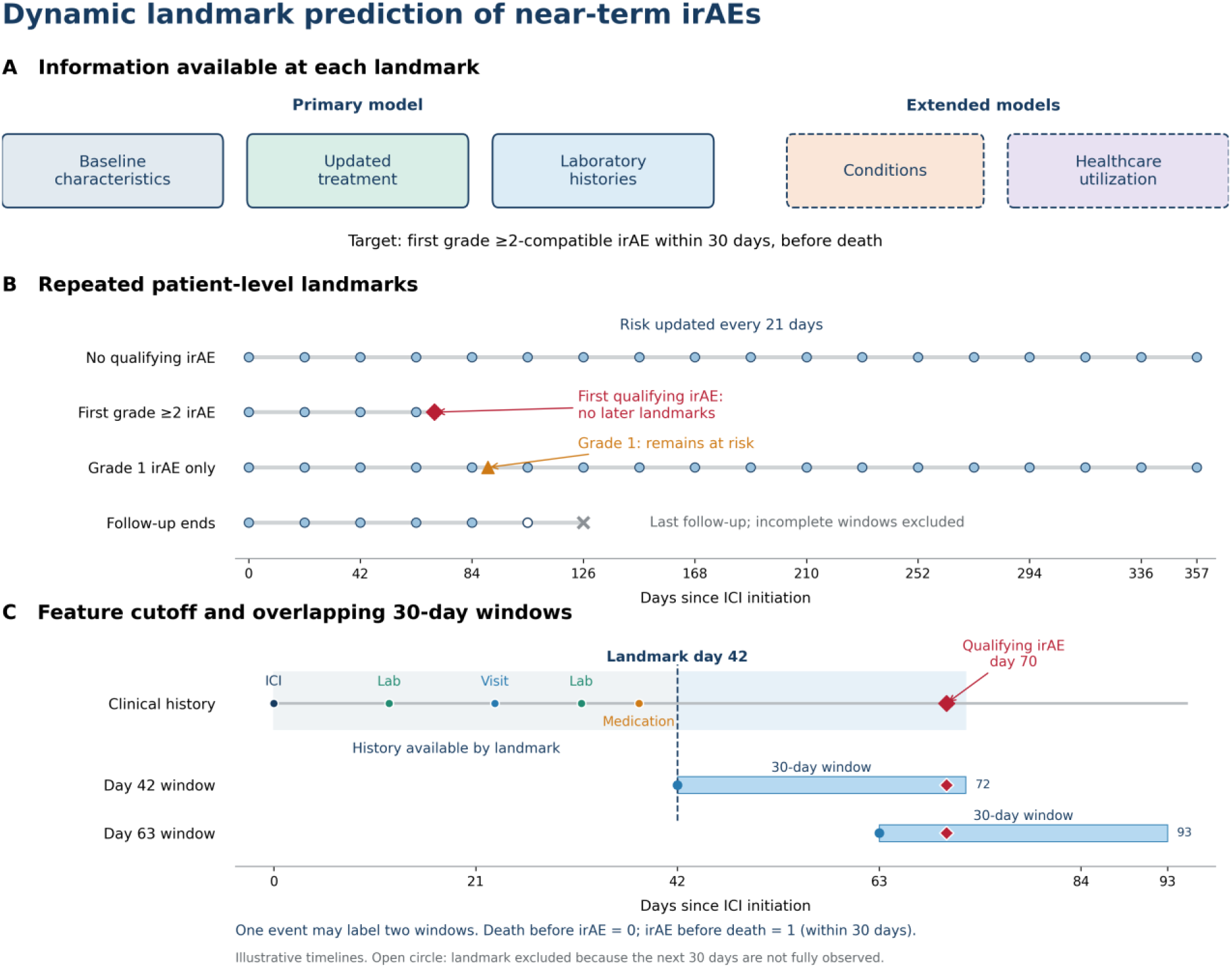
Dynamic landmark design for predicting near-term clinically significant immune-related adverse events (irAEs). (A) The primary model combined baseline characteristics, updated treatment information, and laboratory histories available at each landmark; conditions and healthcare utilization were evaluated in extended models. (B) Patients contributed landmarks every 21 days from ICI initiation through day 357 while alive, under observation, and free of a previous grade ≥2-compatible irAE. Landmarks on or after the first qualifying event were excluded, whereas patients with grade 1 toxicity alone remained at risk. The open circle illustrates a landmark excluded because follow-up ended before completion of its outcome window. (C) Each landmark used information available at or before that date to predict the subsequent 30 days. An illustrative event on day 70 labels both the day 42 and day 63 windows as positive. Death within a prediction window without a preceding qualifying irAE was assigned an outcome of 0; a qualifying irAE occurring within the window before death was assigned an outcome of 1. Patient timelines are illustrative.

## Prediction features

The multiscale feature set contained 218 predictors: 213 numeric and 5 categorical variables. Predictors represented demographics, race and ethnicity, body mass index, cancer group and site, ICI agent and checkpoint target, prior autoimmune disease, comorbidity groups, baseline corticosteroid and chemotherapy exposure, cumulative treatment history, landmark day, and longitudinal laboratory information. Laboratory features included the most recent value, time since measurement, counts of measurements, 30-, 90-, and 180-day means, slopes, changes from baseline, and selected clinically motivated ratios. All predictors were constructed using data available at or before the feature cutoff. Supplementary Table S5 summarizes the predictor families, their temporal construction, and the feature sets evaluated in ablation and extension analyses. Baseline comorbidity concept sets are provided in Supplementary Table S6. Secondary models evaluated additional feature blocks as exploratory analyses. The multiscale trajectory model remained the principal reported model; extensions were interpreted as secondary comparisons rather than selected as definitive winners from temporal performance.

## Model development and comparators

The primary dynamic model used XGBoost gradient-boosted decision trees.^26^ Hyperparameters and boosting duration were selected using patient-level folds within the development cohort and then frozen for feature-ablation and temporal-evaluation analyses. Categorical variables were encoded consistently across datasets, and missing numeric values were handled using XGBoost’s native missing-value support. The principal comparator was a time-aware baseline XGBoost model containing pretreatment information and time since ICI initiation while excluding post-index longitudinal changes. Elastic-net logistic regression was evaluated as a linear comparator using the same multiscale predictors.^27^ Secondary XGBoost models added healthcare-utilization and longitudinal condition-state features to assess their incremental contribution. Because the 2023 cohort was examined during model development, it was described as an internal temporal evaluation cohort rather than an external validation cohort. Model specifications and final settings are provided in Supplementary Table S7.

## Statistical analysis

We evaluated pooled area under the receiver operating characteristic curve (AUROC), pooled area under the precision–recall curve (AUPRC), Brier score, log loss, and mean predicted risk.^28,29^ Because pooled metrics can capture differences in baseline risk across treatment phases, we also calculated AUROC within each landmark and combined landmark-specific AUROCs using the number of positive–negative pairs as weights. Landmark-specific AUPRCs were summarized using row weights. Calibration was assessed by comparing mean predicted risk with observed risk and through graphical calibration analyses.^30^ Incremental performance was estimated using paired patient-clustered bootstrap resampling.^31^ In each replicate, patients rather than individual landmark rows were sampled with replacement, thereby preserving each selected patient’s complete longitudinal record and the dependence among repeated observations. We calculated percentile 95% confidence intervals for each model and for paired differences between the dynamic and baseline models. Primary comparisons used 2,000 bootstrap replicates. Sensitivity analyses varied the prediction horizon, landmark cadence, follow-up requirements, and feature representation. For within-landmark AUROC, only landmarks containing both outcome classes contributed, with weights proportional to the number of positive–negative pairs. Overlapping prediction windows allowed one event to label multiple eligible landmarks; landmark-level event prevalence was therefore distinct from the proportion of unique patients who experienced an event. Reporting followed the data-partition, repeated-observation, outcome-definition, performance-evaluation, and reproducibility recommendations of TRIPOD+AI.^32^

## Results

### Cohort and landmark observations

The development cohort included 1,522 patients contributing 18,799 candidate landmark observations; 495 observations from 350 patients were followed by a grade 2 or higher irAE within 30 days. The 2023 temporal cohort included 526 patients contributing 6,285 candidate landmark observations; 184 observations from 134 patients were positive. No patient appeared in both cohorts. Because prediction windows overlapped, an affected patient could contribute more than one positive landmark. A mutually exclusive follow-up audit identified 55 development and 22 temporal-evaluation landmarks without adequate 30-day outcome ascertainment after accounting for competing death. Excluding these administratively censored observations yielded 18,744 development and 6,263 temporal-evaluation landmarks without changing the number of positive observations. The resulting temporal landmark-level event rate was 2.94%. Patient characteristics at ICI initiation are reported in Table 1; measurable temporal case-mix changes included sex, cancer distribution, corticosteroid exposure, and comorbidity burden.

**Table 1.** Characteristics at ICI initiation

| Characteristic | Development<br>(N=1522) | Temporal<br>evaluation<br>(N=526) | Absolute<br>SMD |
| --- | --- | --- | --- |
| Demographic characteristics |  |  |  |
| Age at ICI initiation, mean (SD), years | 67.0 (13.9) | 67.1 (13.5) | 0.006 |
| Male sex, n (%) | 824 (54.1) | 236 (44.9) | 0.186 |
| Anthropometric characteristics |  |  |  |
| Body mass index, median [IQR], kg/m<sup>2</sup> | 25.7 [22.6-29.5] | 25.6 [22.2-29.7] | 0.029 |
| Cancer group, n (%) |  |  |  |
| Lung | 306 (20.1) | 102 (19.4) | 0.018 |
| GI and hepatobiliary | 277 (18.2) | 89 (16.9) | 0.034 |
| Genitourinary | 235 (15.4) | 74 (14.1) | 0.039 |
| Multiple or unknown | 199 (13.1) | 50 (9.5) | 0.113 |
| Skin and melanoma | 124 (8.1) | 51 (9.7) | 0.054 |
| Gynecologic | 95 (6.2) | 49 (9.3) | 0.115 |
| Breast | 79 (5.2) | 57 (10.8) | 0.209 |
| Other | 70 (4.6) | 12 (2.3) | 0.127 |
| Central nervous system | 64 (4.2) | 14 (2.7) | 0.085 |
| Sarcoma and bone | 43 (2.8) | 10 (1.9) | 0.061 |
| Hematologic | 30 (2.0) | 18 (3.4) | 0.090 |
| Index ICI regimen, n (%) |  |  |  |
| PD-1 monotherapy | 1167 (76.7) | 383 (72.8) | 0.089 |
| PD-L1 monotherapy | 219 (14.4) | 68 (12.9) | 0.043 |
| PD-1 + CTLA-4 | 112 (7.4) | 51 (9.7) | 0.084 |
| PD-1 + LAG-3 | 10 (0.7) | 10 (1.9) | 0.111 |
| CTLA-4 monotherapy | 14 (0.9) | 4 (0.8) | 0.017 |
| Other ICI combination | 0 (0.0) | 10 (1.9) | 0.197 |
| Pretreatment clinical characteristics |  |  |  |
| Prior autoimmune disease, n (%) | 108 (7.1) | 45 (8.6) | 0.054 |
| Baseline systemic corticosteroid exposure, n (%) | 901 (59.2) | 358 (68.1) | 0.185 |
| Prior chemotherapy, n (%) | 578 (38.0) | 171 (32.5) | 0.115 |
| Number of comorbidity groups, median [IQR] | 1 [0-2] | 2 [1-3] | 0.362 |

## Structured phenotype assessment

Chart review was completed for 296 patients sampled across four phenotype-rule groups and a phenotype-negative control group: 60 patients each from Rules 1, 3, and 4 and the negative group, and 56 from Rule 2. Review confirmed grade ≥2 irAEs in 186 patients; 90 were classified as non– grade ≥2 or rejected, and 20 remained indeterminate (Supplementary Table S4). Confirmation varied across sampling rules. Among repeated diagnosis-only candidates (Rule 1), 36 of 60 were confirmed as grade ≥2 (60.0%; 95% CI, 46.5-72.4), increasing to 65.5% among determinate cases. Among diagnosis-plus-systemic-treatment candidates (Rule 2), 47 of 56 were confirmed (83.9%; 95% CI, 71.7-92.4), or 88.7% among determinate cases. Rules 3 and 4 yielded confirmation proportions of 75.0% (45/60; 95% CI, 62.1-85.3) and 96.7% (58/60; 95% CI, 88.5-99.6), respectively; corresponding proportions among determinate cases were 90.0% and 98.3%. No grade ≥2 irAEs were identified among the 60 phenotype-negative controls: 59 were confirmed negative and one was indeterminate. Across all sampling groups, confirmation was 62.8% among all reviewed patients and 67.4% among determinate cases. These aggregate proportions describe the stratified review sample rather than cohort-wide predictive values. The substantial grade ≥2 confirmation among presumed lower-severity candidates indicates that diagnosis-only classification did not reliably distinguish mild from clinically significant toxicity.

## Primary dynamic model performance

In the 2023 temporal evaluation cohort, the multiscale XGBoost model achieved pooled AUROC 0.716 (95% CI, 0.676-0.755), pooled AUPRC 0.067, pair-weighted within-landmark AUROC 0.672 (95% CI, 0.631-0.716), row-weighted landmark AUPRC 0.080, Brier score 0.0281, and log loss 0.1245. The time-aware baseline achieved pooled AUROC 0.631 (95% CI, 0.582-0.679), pooled AUPRC 0.054, within-landmark AUROC 0.536 (95% CI, 0.483-0.592), Brier score 0.0284, and log loss 0.1296. Elastic-net logistic regression achieved AUROC 0.686 and AUPRC 0.058. A complete clinical-state XGBoost extension that also incorporated repeated condition and utilization features achieved AUROC 0.719 and AUPRC 0.073. Calibration curves showed broad agreement between predicted and observed risk across most of the prediction range, with greater variability in the highest-risk bins (Figure 2; Table 2).

**Figure 2.**
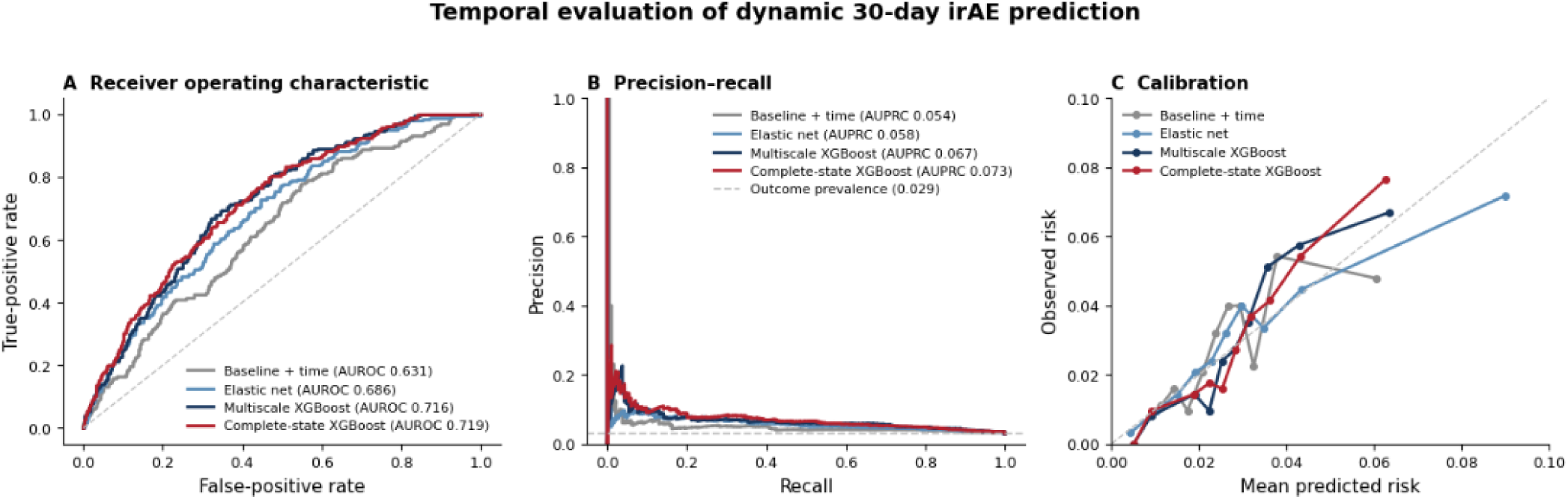
**Internal temporal performance for 30-day irAE prediction**. A, Pooled receiver operating characteristic curves. B, Precision–recall curves, with landmark-level prevalence as the no-skill reference. C, Calibration plots comparing mean predicted and observed risks. Models comprise baseline plus time, elastic net, multiscale XGBoost, and the complete-state extension. The temporal cohort was examined during development.

**Table 2.** Thirty-day prediction performance in the 2023 internal temporal cohort

| Model | Pooled AUROC | Pooled AUPRC | Within-landmark AUROC | Brier score | Log loss |
| --- | --- | --- | --- | --- | --- |
| Baseline plus time | 0.631 (0.582-0.679) | 0.054 | 0.536 (0.483-0.592)* | 0.0284 | 0.1296 |
| Elastic net | 0.686 | 0.058 | — | — | — |
| Multiscale XGBoost | 0.716 (0.676-0.755) | 0.067 | 0.672 (0.631-0.716) | 0.0281 | 0.1245 |
| Complete-state XGBoost | 0.719 | 0.073 | 0.674 | — | — |

\*Values in parentheses are patient-clustered bootstrap 95% confidence intervals. AUROC, area under the receiver operating characteristic curve; AUPRC, area under the precision–recall curve. Dashes indicate estimates not reported. Within-landmark AUROC was weighted by positive-negative pairs.

## Sensitivity and ablation analyses

Sequential feature-state analysis demonstrated increasing discrimination as updated information was incorporated. Pooled AUROC increased from 0.631 for baseline characteristics and landmark time to 0.669 after adding updated ICI exposure, 0.687 after adding the current laboratory state, and 0.716 after incorporating multiscale laboratory trajectories. Corresponding within-landmark AUROCs were 0.536, 0.606, 0.630, and 0.672. Adding recent medication and healthcare-utilization features produced the highest observed discrimination (pooled AUROC, 0.721; within-landmark AUROC, 0.678), but differences relative to the multiscale model were small and paired bootstrap confidence intervals included no difference. Condition-state features did not improve discrimination. The complete clinical-state model modestly improved Brier score and log loss but not AUROC conclusively (Figure 3A-B; Supplementary Table S8).

**Figure 3.**
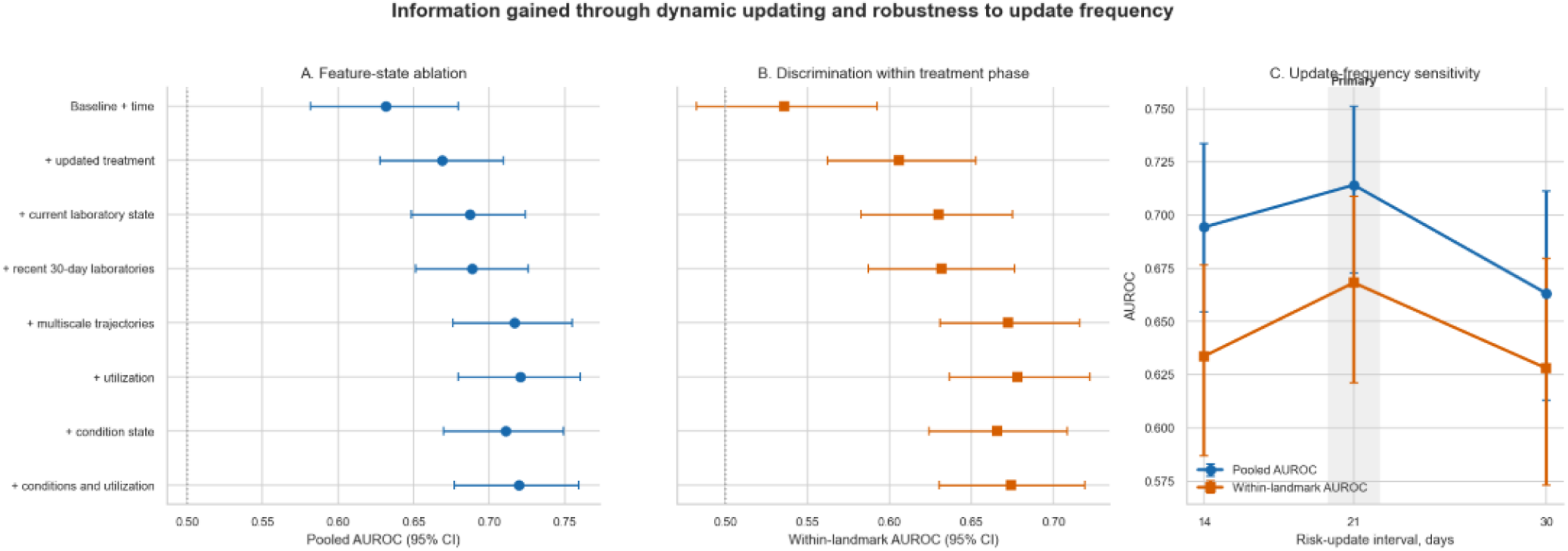
**Feature-state ablations and update-frequency sensitivity**. A, Pooled AUROC for sequential feature states. B, Pair-weighted within-landmark AUROC. C, Discrimination under 14-, 21-, and 30-day updating with a fixed 30-day horizon. Error bars represent patient-clustered bootstrap 95% confidence intervals. The cadence analysis was regenerated separately and yielded slightly different 21-day estimates from the principal feature-state analysis; schedules also differed in prediction opportunities and event prevalence.

Cadence sensitivity analysis showed the highest discrimination with the primary 21-day schedule. With the 30-day prediction horizon held constant, pooled AUROC was 0.694 with 14-day updating, 0.714 with 21-day updating, and 0.663 with 30-day updating; corresponding within-landmark AUROCs were 0.634, 0.668, and 0.628. Relative to 21-day updating, the paired pooled-AUROC difference was -0.020 (95% CI, -0.043 to 0.004) for 14-day updating and -0.051 (95% CI, -0.089 to -0.016) for 30-day updating. The paired within-landmark difference was -0.035 (95% CI, -0.061 to -0.006) for 14-day updating and -0.040 (95% CI, -0.086 to 0.002) for 30-day updating. AUPRC and raw loss measures were interpreted in the context of differing landmark-level event prevalence across schedules (Figure 3C; Supplementary Table S9).

Near-term prediction was more discriminative than longer-horizon prediction: pooled AUROC was 0.716 for 30 days, 0.686 for 60 days, and 0.679 for 90 days (Supplementary Table S10). More complex architectures did not consistently improve performance. Pairwise ranking achieved pooled AUROC 0.619, and a late-fusion model combining static, treatment, laboratory, immune, and full-dynamic component predictions achieved AUROC 0.708. Multiorgan laboratory perturbation, explicit risk-memory features, a broad diagnosis-prodrome block, and engineered PNI, LMR, SII, SIRI, AISI, and LDH-to-albumin features produced small, inconsistent, or adverse changes and were not retained in the primary representation (Supplementary Table S11).

## Discussion

In this pan-cancer EHR cohort, repeated updating of clinical information improved prediction of a first grade ≥2-compatible irAE within the subsequent 30 days. The multiscale trajectory model achieved pooled AUROC 0.716 and within-landmark AUROC 0.672, compared with 0.631 and 0.536 for pretreatment characteristics plus treatment time. Updated ICI exposure and laboratory trajectories accounted for most of the improvement. Utilization provided a smaller increment whose paired confidence intervals included no difference, while repeated condition features did not improve discrimination. Previous studies offer complementary approaches: Iivanainen et al. reported AUROC 0.93 for irAE onset using repeated electronic patient-reported symptoms in 34 patients,^19^ and He et al. reported AUROC 0.902 using a heterogeneous EHR graph model for patient-level irAE classification.^18^ Early B-cell changes, immune signatures, and pretreatment tissue biomarkers have also been associated with subsequent toxicity.^15,33,34^ These studies support toxicity risk assessment using diverse information sources, but differences in populations, severity definitions, prediction timing, and validation design prevent direct comparisons with our fixed-horizon task.

The aggregate endpoint spans cancer types, regimens, and organ systems with different mechanisms and precursor signals. Patients with grade 1 toxicity alone remained at risk, and each prediction concerned a new grade ≥2-compatible event within 30 days. Temporal landmark prevalence was 2.94%, giving a no-skill precision–recall reference of approximately 0.029.^29^ Pooled AUPRC values of 0.067 for the multiscale model and 0.073 for the complete-state extension were approximately 2.3 and 2.5 times this reference. This enrichment describes ranking across prediction opportunities rather than the positive predictive value of a chosen alert threshold. Within-landmark improvement further indicates that dynamic information distinguished patients evaluated at comparable treatment times, beyond differences between early and late treatment periods. Brier score and log loss improved modestly, while calibration plots showed greater uncertainty at higher predicted risks; calibration intercept, slope, and threshold-specific alert performance require further evaluation.^30^ The 21-day cadence had the highest observed discrimination, but cadence changes also alter prediction opportunities, overlapping windows, and prevalence. These findings support the feasibility of three-week updating without establishing a biologically optimal interval.

Chart review highlighted both the usefulness and limitations of structured ascertainment. Among 296 reviewed patients, 186 had confirmed grade ≥2 irAEs, 90 were classified as non–grade ≥2 or rejected, and 20 remained indeterminate. Confirmation varied substantially across rule groups. Notably, 36 of 60 presumed lower-severity, diagnosis-only candidates were confirmed as grade ≥2, indicating that absence of a structured treatment anchor did not reliably establish mild toxicity. Among 60 phenotype-negative controls, 59 were confirmed negative and one was indeterminate. Because sampling was stratified by rule, aggregate confirmation proportions do not represent cohort-wide predictive values. Attribution and severity remain difficult when infection, progression, other treatment effects, or preexisting disease resemble irAEs. Onset-date discrepancies can also shift events across prediction windows. These findings support further refinement of severity rules and sensitivity analyses using adjudicated status and dates. Narrative phenotyping systems, including irAE-GPT and agentic approaches, may improve extraction of documented toxicity states, although accurate detection alone does not establish future-risk prediction.^6,8^

Strengths include repeated assessment across heterogeneous patients, explicit event-free risk sets, restriction of predictors to available information, and patient-level resampling. Limitations include the single-system setting and examination of the 2023 cohort during development, which may yield optimistic internal temporal performance. Requiring two ICI exposure dates may exclude patients who discontinue after an early severe event. Laboratory frequency and utilization may reflect clinician concern or emerging symptoms as well as biology, making feature-lag analyses and external evaluation important. Overlapping windows do not create independent events, and rare organ-specific toxicities may be poorly represented by an aggregate endpoint. Future studies should incorporate adjudicated onset dates, grade 1 states, and organ-specific narrative signals, then evaluate external calibration, subgroup performance, lead time, and decision-curve net benefit.^35^ Prospective silent evaluation should determine whether risk-guided surveillance detects toxicity earlier at an acceptable alert burden. The present findings support further development of adaptive monitoring but do not establish that model-guided treatment interruption, immunosuppression, or rechallenge improves outcomes.

## Conclusion

A longitudinal EHR model updated every 21 days improved prediction of a first grade ≥2 irAE within 30 days relative to pretreatment characteristics and treatment time. Updated treatment exposure and multiscale laboratory trajectories accounted for most of the gain, including discrimination among patients assessed at comparable treatment times. These findings support further development of adaptive toxicity surveillance, with adjudicated outcomes and independent evaluation required before clinical use.

## Data Availability

The patient-level electronic health record data used in this study are not publicly available because of patient privacy and institutional access restrictions. Aggregate results are provided in the manuscript and supplementary materia

## Acknowledgments

The authors thank the Columbia University Herbert Irving Comprehensive Cancer Center for its support. The authors thank Benjamin May and Jianhua Liu for curating the data.

## Author contributions

Liyuan Gong (Conceptualization, Data curation, Formal analysis, Investigation, Methodology, Project administration, Supervision, Validation, Visualization, Writing-original draft, Writing-review & editing), Alice Agyekum (Formal analysis, Validation, Writing-review & editing), Nitish Aswani (Validation, Writing-review & editing), Harry Reyes Nieva (Conceptualization, Investigation, Methodology, Writing-review & editing), and Chin Hur (Funding acquisition, Supervision, Validation, Writing-review & editing)

## Data and code availability

The CUMC institutional EHR data used in this study cannot be publicly shared because they contain protected health information and are subject to HIPAA regulations, institutional policies, and data-use restrictions. To support reproducibility while maintaining compliance, the analytic code has been cleaned to remove any institution-specific paths, credentials, identifiers, or protected health information and is available at: https://github.com/abundanceli.

## Artificial intelligence use disclosure

Generative artificial intelligence tools were used to assist with language editing and manuscript organization. All study design decisions, analyses, interpretation, source verification, and final text were reviewed and approved by the authors. No patient-level protected health information was entered into publicly accessible generative artificial intelligence services.

---

**Supplementary Table S1.** ICI drug concept sets defined using RxNorm ingredient concepts and descendant drug concepts.

| **Target** | **Drug name** | **Brand name** | **Ingredient**<br>**Concept Name** | **RxNorm**<br>**Ingredient**<br>**Concept ID** | **Number of**<br>**Descendant**<br>**Concepts** |
| --- | --- | --- | --- | --- | --- |
| Anti-PD-1 | Pembrolizumab | Keytruda | pembrolizumab | 45775965 | 113 |
|  | Nivolumab | Opdivo | nivolumab | 45892628 | 179 |
|  | Cemiplimab | Libtayo | cemiplimab | 35200783 | 32 |
|  | Dostarlimab | Jemperli | dostarlimab | 1536789 | 32 |
| Anti-PD-L1 | Atezolizumab | Tecentriq | atezolizumab | 42629079 | 76 |
|  | Durvalumab | Imfinzi | durvalumab | 1593273 | 44 |
|  | Avelumab | Bavencio | avelumab | 1594034 | 48 |
| Anti-CTLA-4 | Ipilimumab | Yervoy | ipilimumab | 40238188 | 85 |
|  | Tremelimumab | Imjudo | tremelimumab | 741851 | 13 |
| Anti-LAG-3 | Relatlimab | Opdualag | relatlimab | 779239 | 11 |

**Supplementary Table S2.** International Classification of Diseases Diagnosis Codes for irAE.

| Category | Condition | ICD-10 Codes | ICD-9 Codes | Exclude if Pre-existing |
| --- | --- | --- | --- | --- |
| **Dermatologic** | Bullous dermatitis | L13.9 | 694.9 | No |
|  | Drug-related dermatitis / eruption | L27, L27.0–L27.9, L30.9 | 693.0–693.9, 692.9, 995.2 | No |
|  | Pruritus | L29, L29.8–L29.9 | 698.8–698.9 | No |
|  | Erythema multiforme / SJS / TEN | L51.0–L51.9 | 695.10–695.15 | No |
|  | Vitiligo | L80 | 709.01 | No |
|  | Rash | R21 | 782.1 | No |
| **Endocrine** | Hypothyroidism (drug-induced) | E03.2 | 244.3 | No |
|  | Other hypothyroidism | E03.8–E03.9 | 244.8–244.9 | Yes |
|  | Thyrotoxicosis | E05.80–E05.90 | 242.80–242.90 | Yes |
|  | Hypopituitarism | E23.0–E23.7 | 253.2–253.9, 628.1 | Yes |
|  | Adrenal insufficiency | E27.1–E27.4 | 255.4–255.9 | Yes |
| **Gastrointestinal** | Gastritis / duodenitis | K29.0–K29.8 | 535.00–535.60 | No |
|  | Gastroenteritis / colitis | K52.1, K52.8–K52.9 | 558.2–558.9 | No |
|  | Diarrhea | R19.7 | 787.91 | No |
| **Hepatic** | Toxic liver disease | K71 | — | Yes |
|  | Hepatic failure | K72.0–K72.9 | 570, 572.2, 572.8 | Yes |
|  | Autoimmune / inflammatory hepatitis | K75.4, K75.9 | 571.42, 573.3 | Yes |
| **Neurologic** | Myasthenia gravis | G70.00–G70.01 | 358.00–358.01 | Yes |
|  | Encephalitis / myelitis | G04.81–G04.91 | 323.81–323.9 | No |
|  | PRES | I67.83 | 348.39 | No |
| **Pancreatic** | Pancreatitis | K85.3–K85.9 | 577.0 | No |
| **Pulmonary** | Drug-induced<br>interstitial lung disease | J70.2–J70.4 | — | No |
|  | Pneumonitis | J70.8 | 508.8, 486 | No |
| **Renal** | Nephritic syndrome /<br>nephritis | N05.2–N05.9 | 583.0–583.9 | Yes |
|  | Other kidney disorders | N28.8–N28.9 | 593.9 | Yes |
| **Cardiac** | Myocarditis | I40.0–I40.9,<br>I51.4 | 422.0–422.9 | No |

**Supplementary Table S3.**
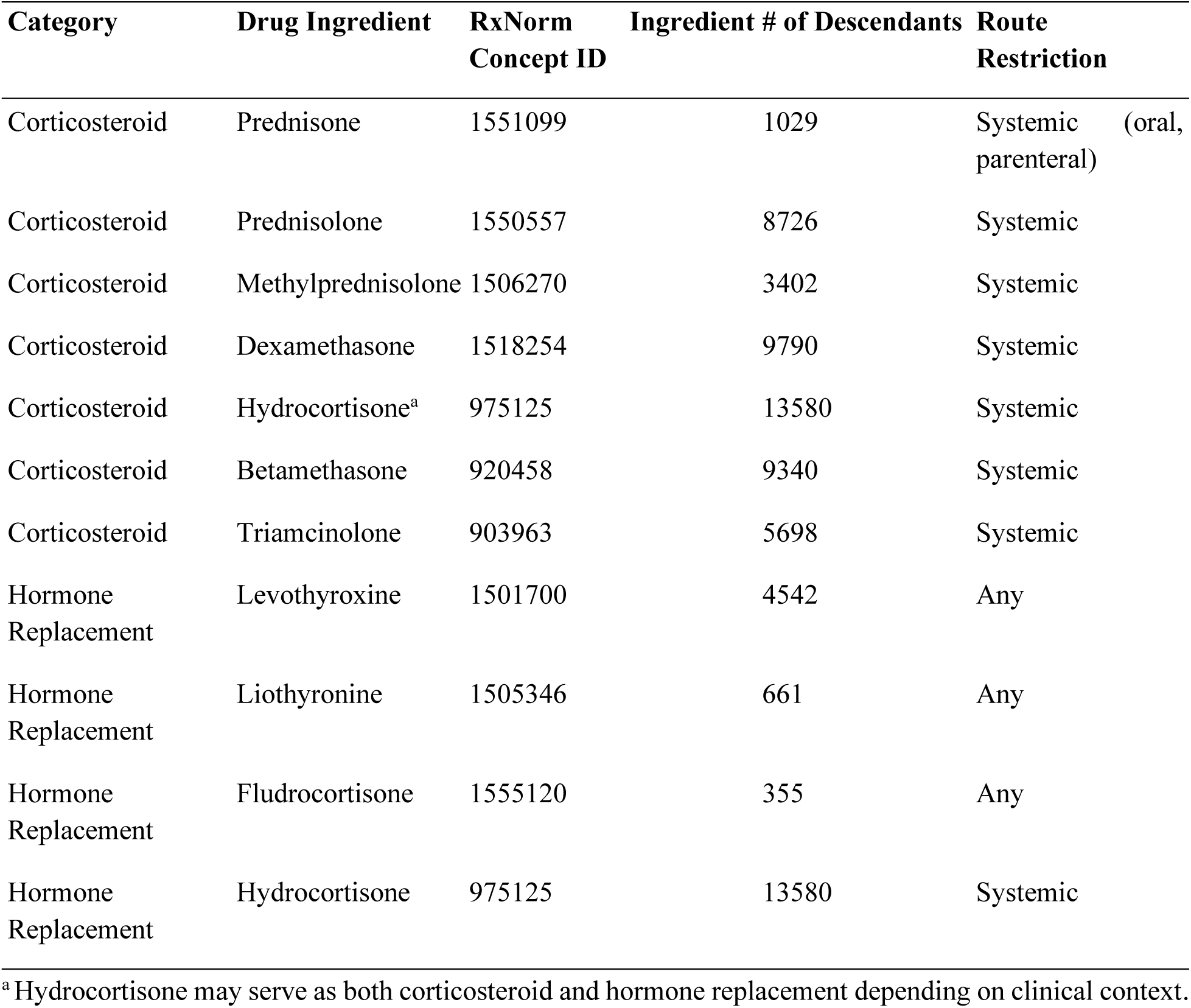
Medication concept sets for treatment-based anchoring of irAEs

| **Category** | **Drug Ingredient** | **RxNorm**<br>**Concept ID** | **Ingredient # of Descendants** | **Route**<br>**Restriction** |
| --- | --- | --- | --- | --- |
| Corticosteroid | Prednisone | 1551099 | 1029 | Systemic (oral,<br>parenteral) |
| Corticosteroid | Prednisolone | 1550557 | 8726 | Systemic |
| Corticosteroid | Methylprednisolone | 1506270 | 3402 | Systemic |
| Corticosteroid | Dexamethasone | 1518254 | 9790 | Systemic |
| Corticosteroid | Hydrocortisone<sup>a</sup> | 975125 | 13580 | Systemic |
| Corticosteroid | Betamethasone | 920458 | 9340 | Systemic |
| Corticosteroid | Triamcinolone | 903963 | 5698 | Systemic |
| Hormone<br>Replacement | Levothyroxine | 1501700 | 4542 | Any |
| Hormone<br>Replacement | Liothyronine | 1505346 | 661 | Any |
| Hormone<br>Replacement | Fludrocortisone | 1555120 | 355 | Any |
| Hormone<br>Replacement | Hydrocortisone | 975125 | 13580 | Systemic |

<sup>a</sup> Hydrocortisone may serve as both corticosteroid and hormone replacement depending on clinical context.

**Supplementary Table S4.** Comorbidity concept sets used for baseline feature extraction

| **Comorbidity Category** | **Condition** | **SNOMED Concept ID** | **Number of Descendant Concepts** |
| --- | --- | --- | --- |
| Chronic lung disease | Chronic obstructive pulmonary disease | 255573 | 45 |
| Chronic lung disease | Asthma | 317009 | 113 |
| Chronic lung disease | Interstitial lung disease | 4119786 | 72 |
| Cardiovascular | Coronary arteriosclerosis | 317576 | 43 |
| Cardiovascular | Myocardial infarction | 4329847 | 112 |
| Diabetes | Type 1 diabetes mellitus | 201254 | 17 |
| Diabetes | Type 2 diabetes mellitus | 201826 | 18 |
| Thyroid disease | Hypothyroidism | 140673 | 101 |
| Thyroid disease | Hyperthyroidism | 4142479 | 22 |
| Thyroid disease | Thyroiditis | 133444 | 137 |
| Chronic liver disease | Chronic hepatitis | 200763 | 34 |
| Chronic liver disease | Cirrhosis of liver | 4064161 | 62 |
| Chronic liver disease | Non-alcoholic fatty liver | 4026131 | 2 |
| Chronic Kidney disease | Chronic kidney disease | 46271022 | 143 |
| Autoimmune | Psoriasis | 140168 | 90 |
| Autoimmune | Systemic lupus erythematosus | 257628 | 35 |
| Autoimmune | Rheumatoid arthritis | 80809 | 145 |
| Autoimmune | Inflammatory bowel disease | 4074815 | 23 |

**Supplementary Table S4.**
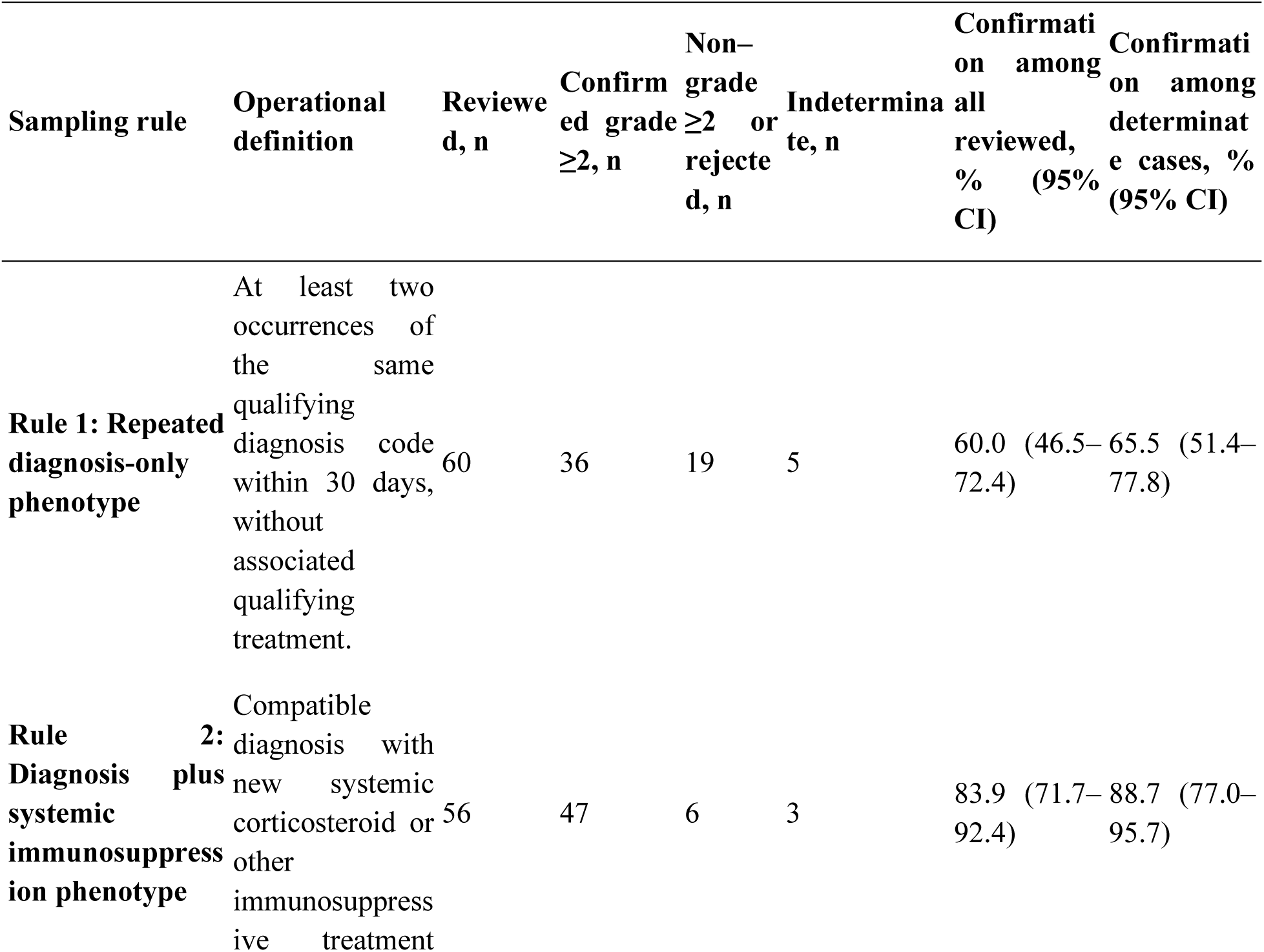
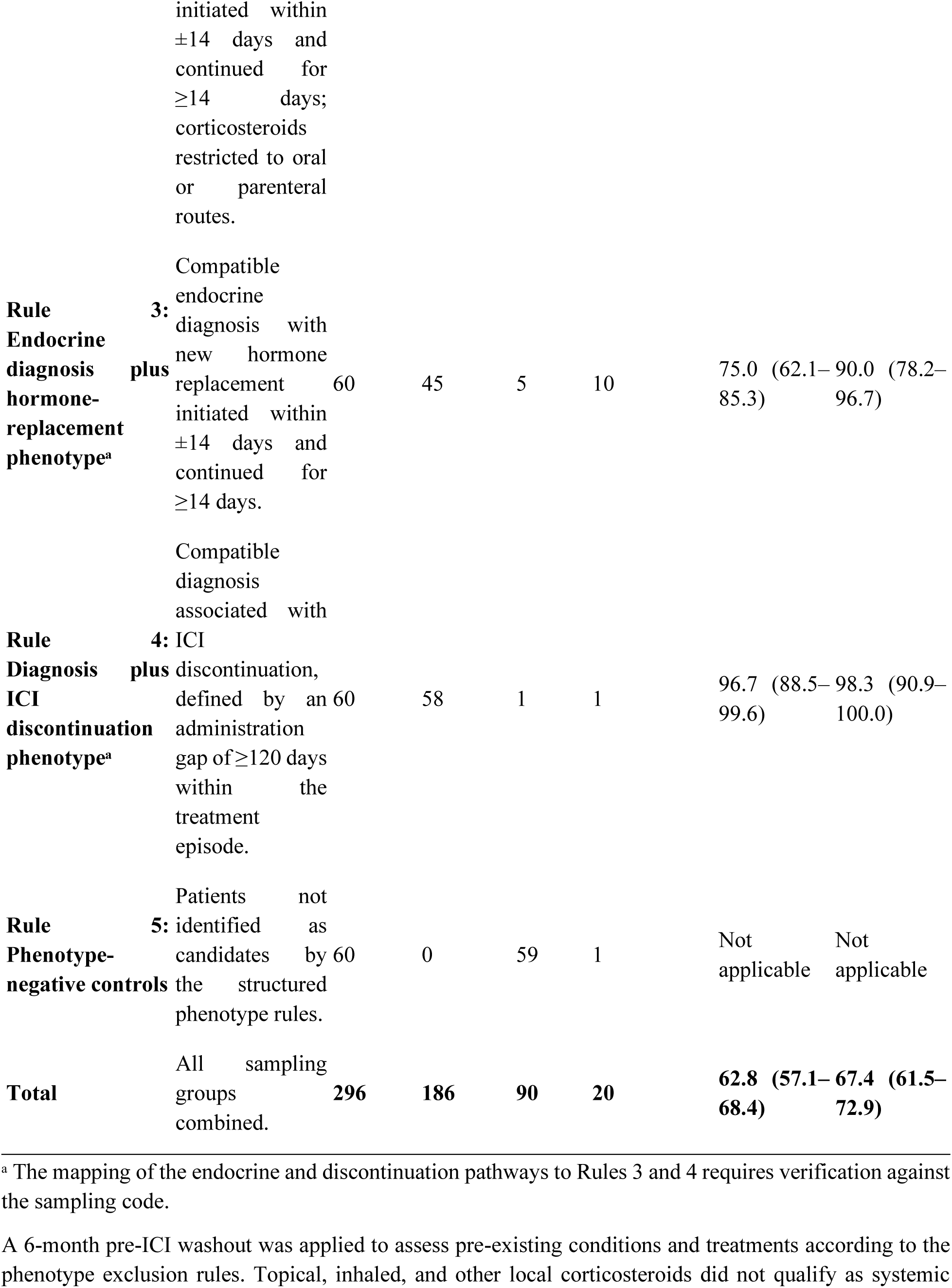
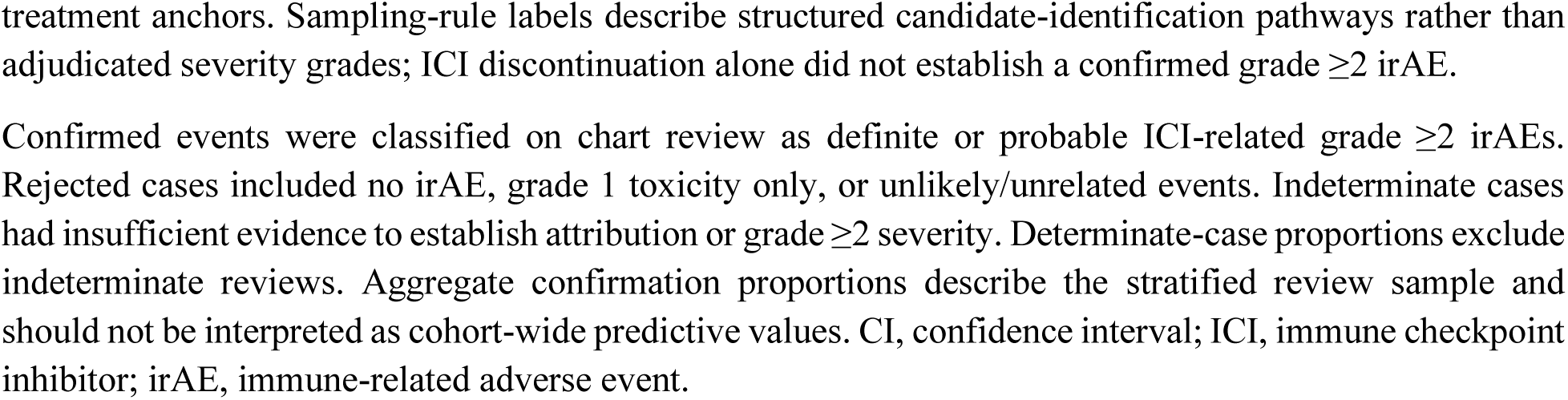
Chart-review findings by structured phenotype sampling rule

**Supplementary Table S5.** Prediction-feature families S5A. Primary multiscale predictors. The four ratios in this classification are dyn_NLR, dyn_PLR, dyn_SII, and dyn_CRP_albumin_ratio. dyn_MLR was classified in the existing 58-variable nonlaboratory/other group. We can either preserve that exact computational grouping or move MLR into the ratio family and change the displayed counts to 5 and 57. Preserving the original grouping is preferable for exact reproducibility.

| Feature family | Predictors, n | Information represented | Temporal construction |
| --- | --- | --- | --- |
| Nonlaboratory and other predictors | 58 | Landmark day, demographics, race and ethnicity, BMI, cancer group and site, baseline ICI regimen, autoimmune history, comorbidities, baseline corticosteroid and chemotherapy exposure, baseline laboratory summaries, and updated ICI-treatment history | Static variables fixed at ICI initiation; treatment variables updated through the landmark cutoff |
| Current laboratory values | 27 | Baseline and most recently available values; deviations of the latest value from recent averages | Latest result available at or before the feature cutoff |
| Laboratory recency | 21 | Time since each laboratory measurement or derived ratio and time since the last ICI administration | Days between the landmark cutoff and most recent qualifying measurement or exposure |
| Laboratory 30-day summaries | 27 | Means during the previous 30 days and contrasts between recent and longer-term averages | Rolling 30-day window ending at the feature cutoff |
| Laboratory 90-day summaries | 15 | Mean laboratory values during the previous 90 days | Rolling 90-day window ending at the feature cutoff |
| Laboratory 180-day summaries | 15 | Mean laboratory values during the previous 180 days | Rolling 180-day window ending at the feature cutoff |
| Laboratory slopes | 21 | Direction and rate of laboratory change | Linear slope estimated from qualifying measurements during the previous 180 days |
| Change from baseline | 15 | Difference between the most recent value and the pretreatment reference value | Latest value minus baseline value |
| Measurement frequency | 15 | Number of measurements obtained during the previous 180 days | Count through the feature cutoff |
| Derived laboratory ratios | 4 | Neutrophil-to-lymphocyte ratio, platelet-to-lymphocyte ratio, systemic immune- inflammation index, and C-reactive protein-to-albumin ratio | Calculated from the most recent jointly available components |
| **Total** | **218** |  |  |

S5B. Cumulative ablation feature states
| Feature state | Total predictors | Newly added predictors | Purpose |
| --- | --- | --- | --- |
| Baseline + time | 48 | 48 | Pretreatment clinical state plus landmark day |
| + updated treatment | 70 | 22 | Cumulative and recent ICI exposure |
| + current laboratory state | 110 | 40 | Latest values, laboratory recency, and selected ratios |
| + recent 30-day laboratory state | 131 | 21 | Recent means and short-term deviations |
| + multiscale trajectories | 218 | 87 | Longer-term means, slopes, counts, and baseline changes |
| + healthcare utilization | 235 | 17 | Recent outpatient, emergency, inpatient, and medication utilization |
| + condition state | 246 | 28 | Repeated diagnosis/prodrome state |
| + conditions and utilization | 263 | 45 | Complete exploratory EHR state |

**Supplementary Table S6.**
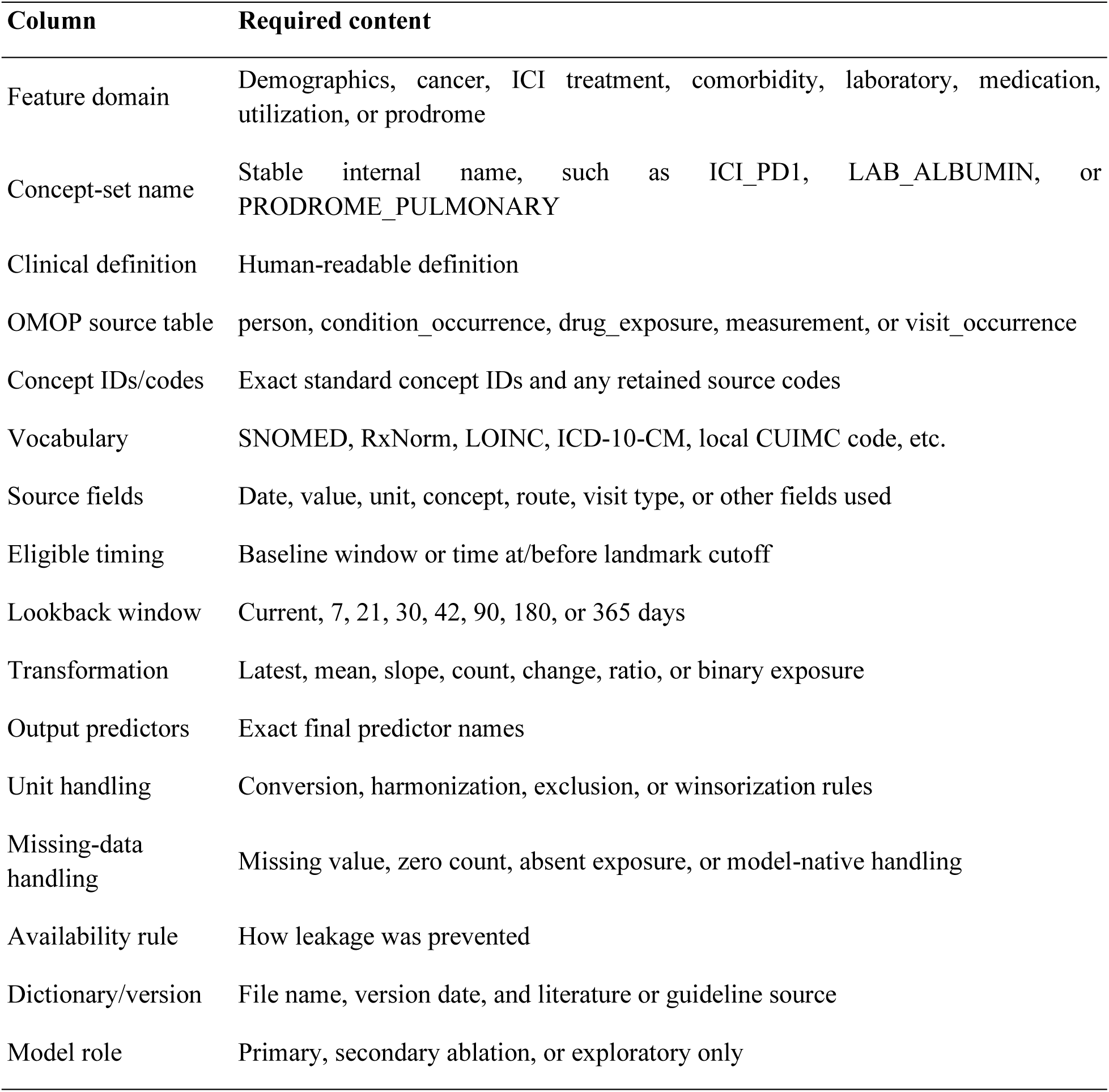
Recommended structure. S6 should have one row per source construct or concept set-not one row per patient and not necessarily one row per final predictor.

| Column | Required content |
| --- | --- |
| Feature domain | Demographics, cancer, ICI treatment, comorbidity, laboratory, medication, utilization, or prodrome |
| Concept-set name | Stable internal name, such as ICI_PD1, LAB_ALBUMIN, or PRODROME_PULMONARY |
| Clinical definition | Human-readable definition |
| OMOP source table | person, condition_occurrence, drug_exposure, measurement, or visit_occurrence |
| Concept IDs/codes | Exact standard concept IDs and any retained source codes |
| Vocabulary | SNOMED, RxNorm, LOINC, ICD-10-CM, local CUIMC code, etc. |
| Source fields | Date, value, unit, concept, route, visit type, or other fields used |
| Eligible timing | Baseline window or time at/before landmark cutoff |
| Lookback window | Current, 7, 21, 30, 42, 90, 180, or 365 days |
| Transformation | Latest, mean, slope, count, change, ratio, or binary exposure |
| Output predictors | Exact final predictor names |
| Unit handling | Conversion, harmonization, exclusion, or winsorization rules |
| Missing-data handling | Missing value, zero count, absent exposure, or model-native handling |
| Availability rule | How leakage was prevented |
| Dictionary/version | File name, version date, and literature or guideline source |
| Model role | Primary, secondary ablation, or exploratory only |

**Supplementary Table S7.**
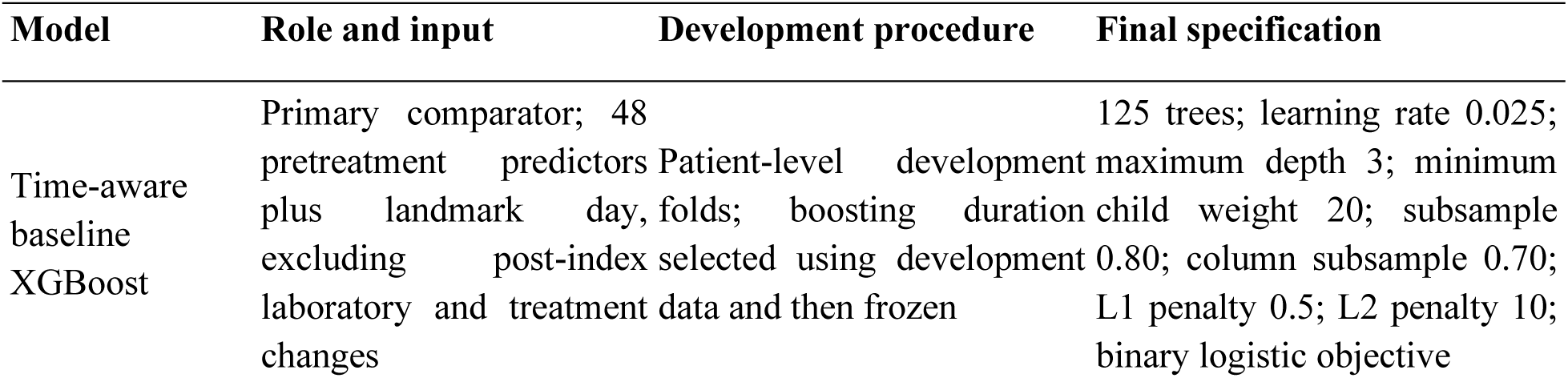
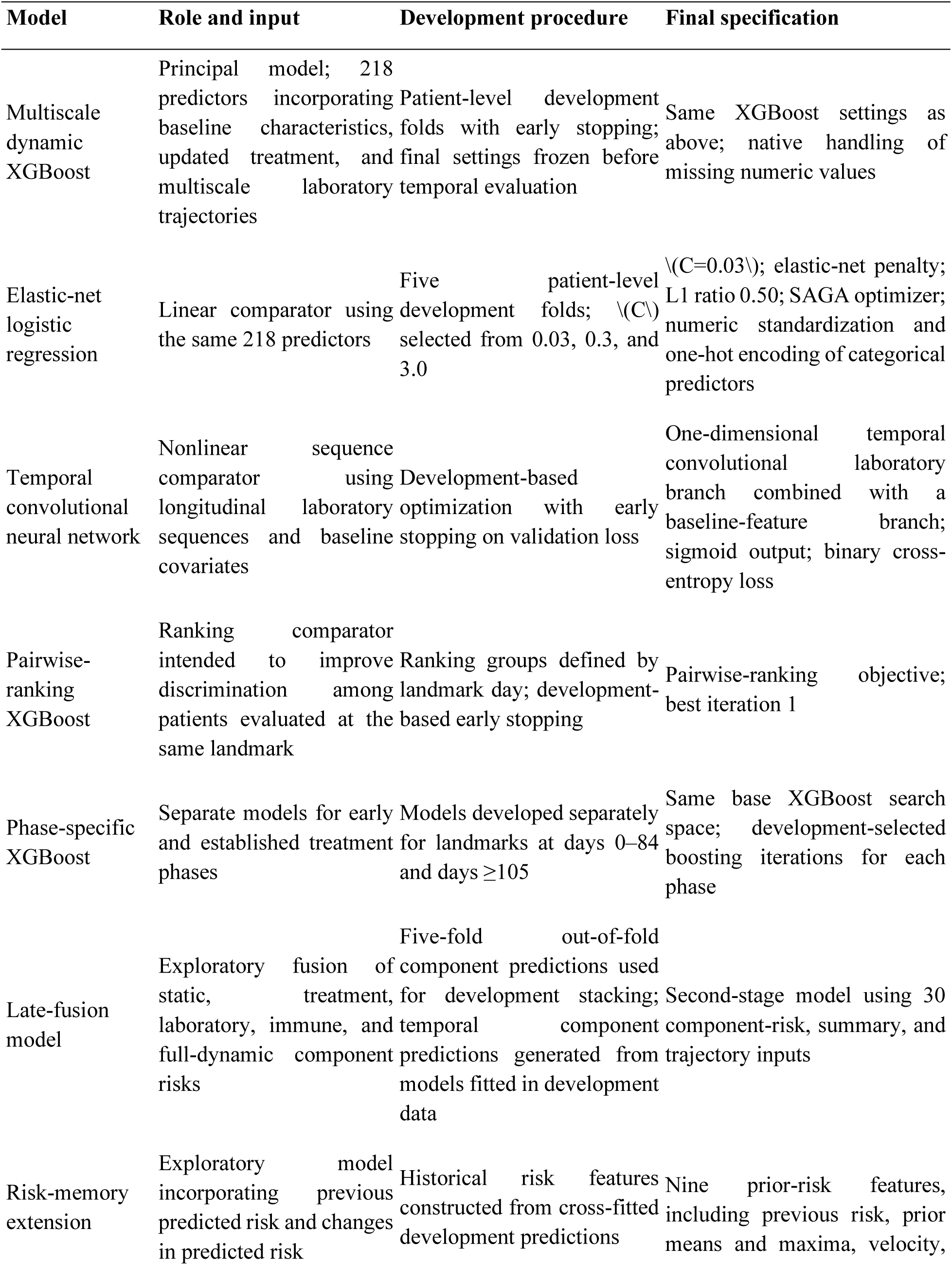
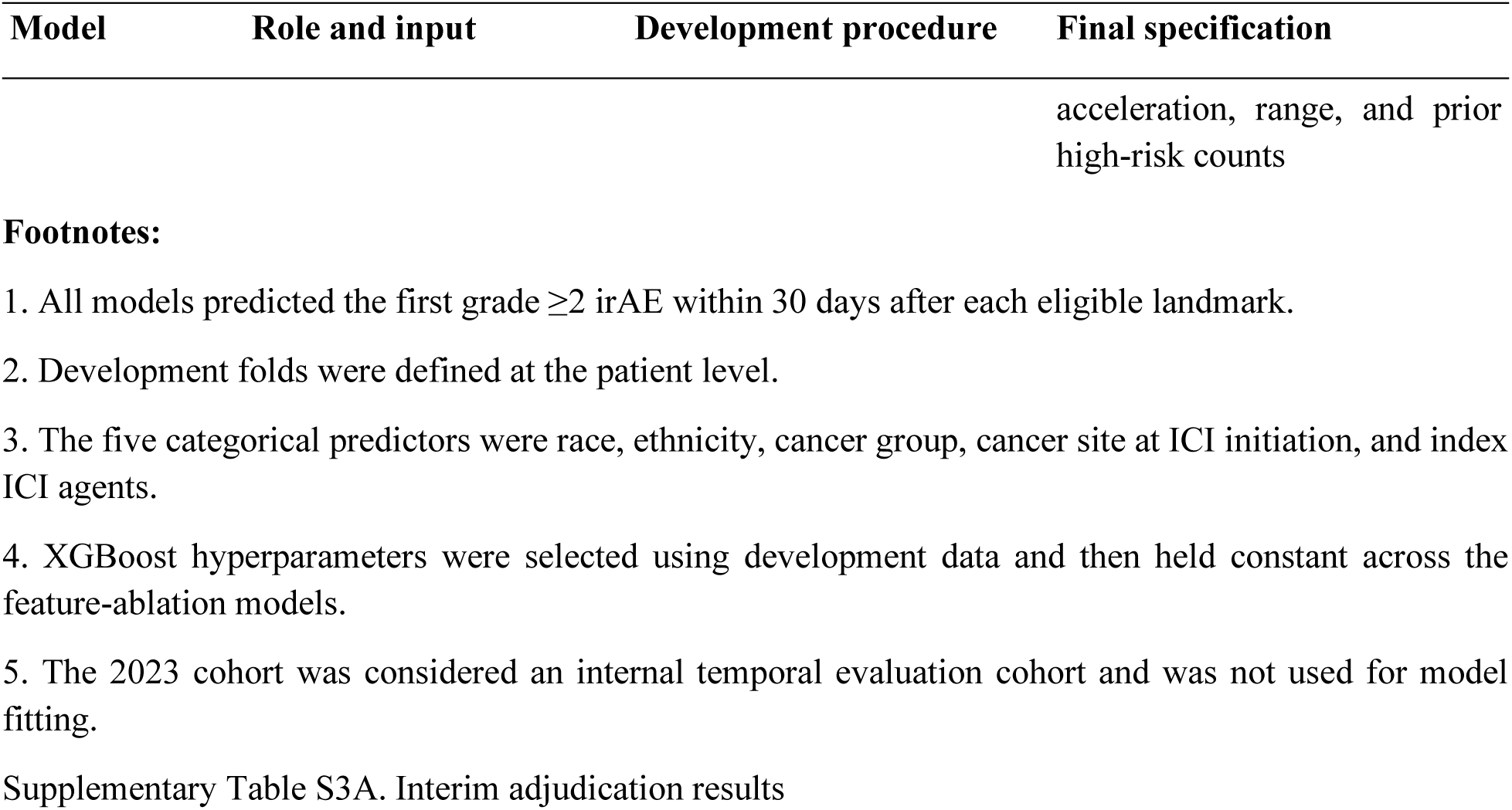
Model specifications and final hyperparameters

**Supplementary Table S8.**
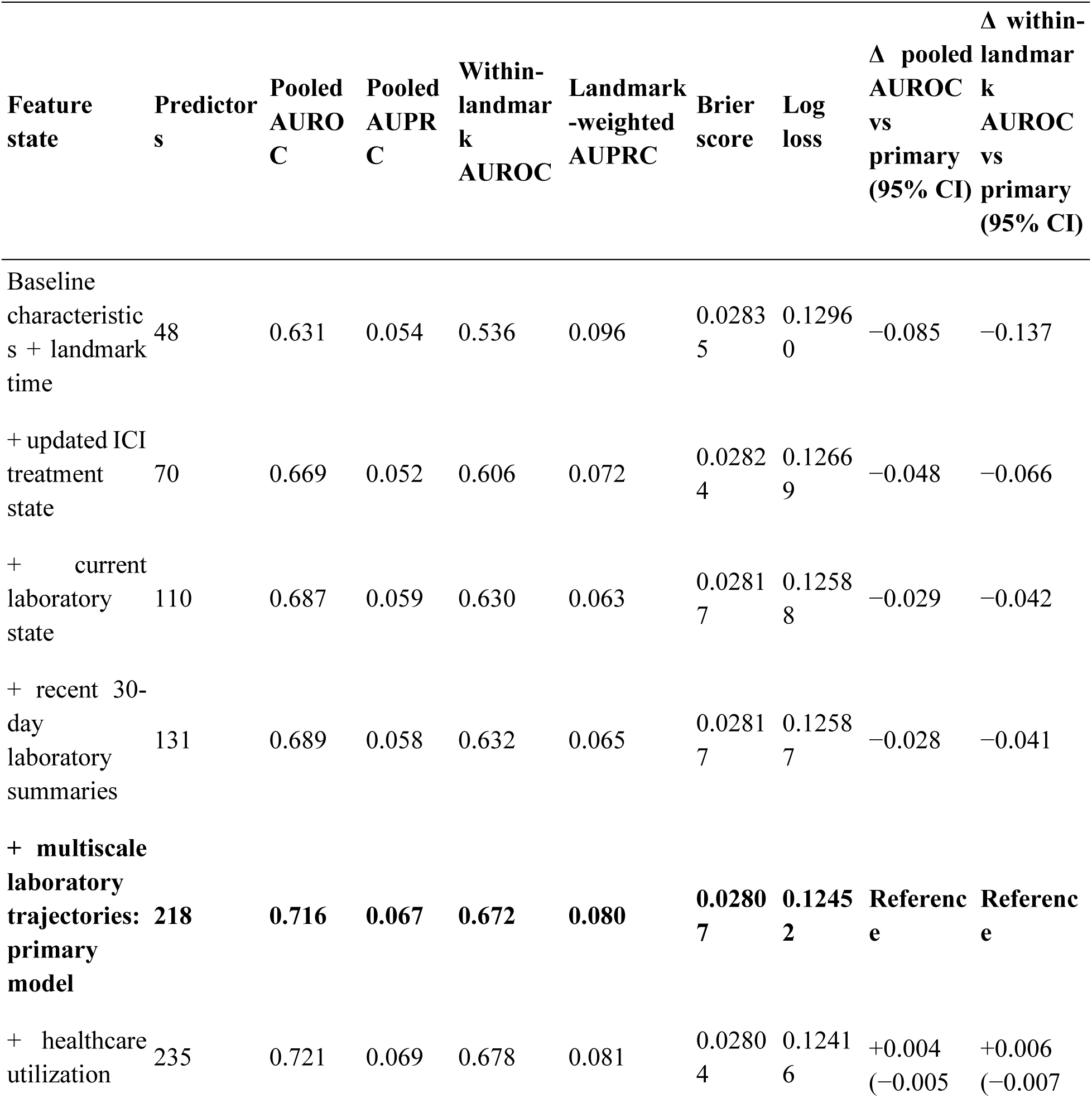
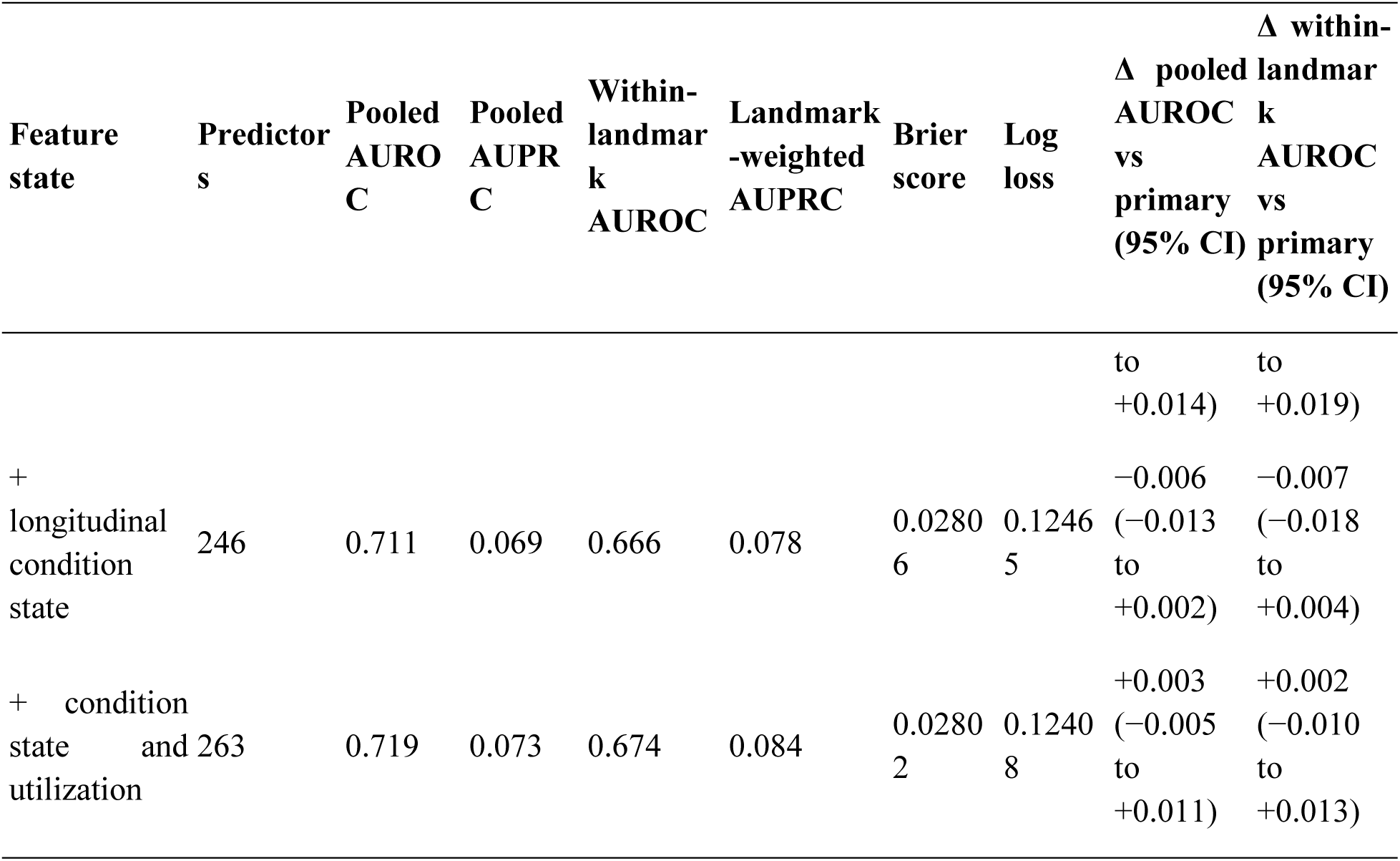
Feature-state ablation and extension performance

**Supplementary Table S9A.**
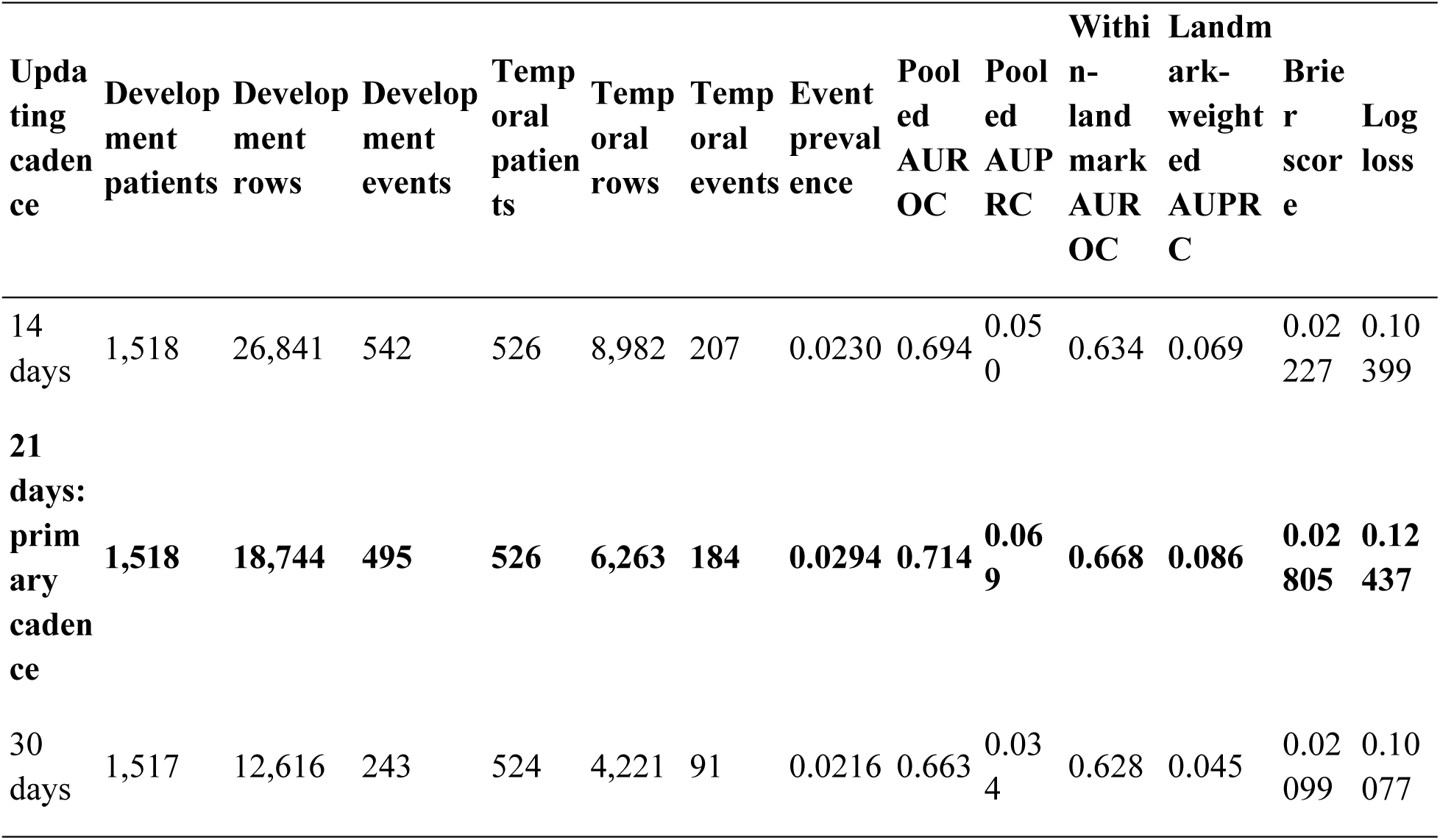
Performance by updating cadence

**Supplementary Table S9B.** Paired differences relative to 21-day updating

S9B. Paired differences relative to 21-day updating
| Comparison | Metric | Difference | 95% CI |
| --- | --- | --- | --- |
| 14-day minus 21-day | Pooled AUROC | -0.0199 | -0.0428 to +0.0036 |
|  | Pooled AUPRC | -0.0192 | -0.0346 to -0.0099 |
|  | Within-landmark AUROC | -0.0345 | -0.0608 to -0.0063 |
|  | Landmark-weighted AUPRC | -0.0173 | -0.0513 to +0.0125 |
|  | Brier score | -0.00579 | -0.00722 to -0.00437 |
|  | Log loss | -0.02039 | -0.02501 to -0.01582 |
| 30-day minus 21-day | Pooled AUROC | -0.0510 | -0.0894 to -0.0163 |
|  | Pooled AUPRC | -0.0347 | -0.0572 to -0.0195 |
|  | Within-landmark AUROC | -0.0402 | -0.0857 to +0.0020 |
|  | Landmark-weighted AUPRC | -0.0405 | -0.0796 to -0.0240 |
|  | Brier score | -0.00707 | -0.00997 to -0.00439 |
|  | Log loss | -0.02363 | -0.03346 to -0.01438 |

**Supplementary Table S10.**
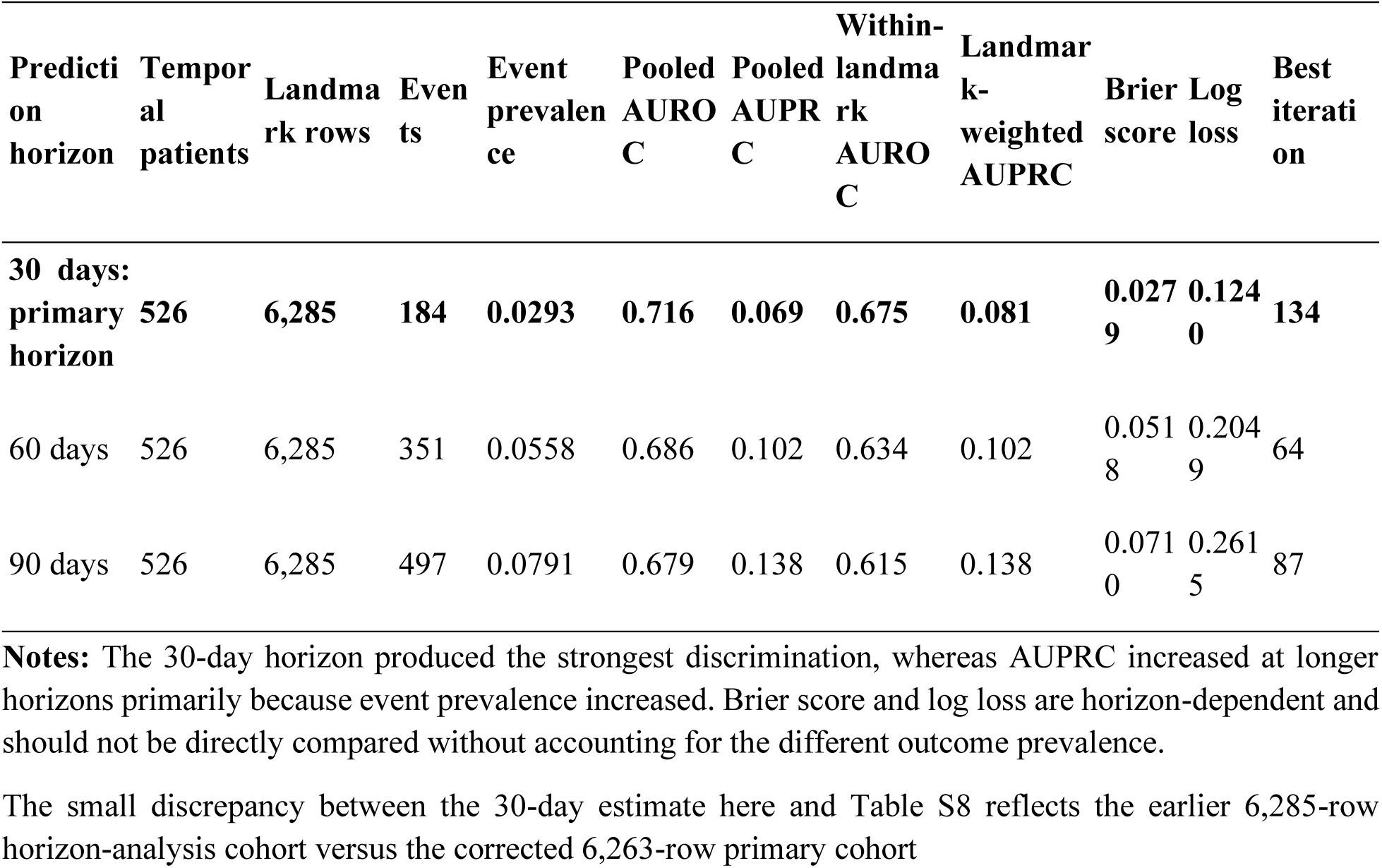
Prediction-horizon sensitivity analysis

